# Myocardial injury is associated with in-hospital mortality of confirmed or suspected COVID-19 in Wuhan, China: A single center retrospective cohort study

**DOI:** 10.1101/2020.03.21.20040121

**Authors:** Fan Zhang, Deyan Yang, Jing Li, Peng Gao, Taibo Chen, Zhongwei Cheng, Kangan Cheng, Quan Fang, Wan Pan, Chunfeng Yi, Hongru Fan, Yonghong Wu, Liwei Li, Yong Fang, Juan Liu, Guowei Tian, Liqun He

**Affiliations:** Department of Cardiology, Wuhan No.1 Hospital; Department of Cardiology, Peking Union Medical College Hospital, Chinese Academy Medical Sciences & Peking Union Medical college, Beijing, China

**Author notes:** Corresponding Author: Liqun He. First Author: Fan Zhang, Co-First Author: Deyan Yang, Fan Zhang and Deyan Yang contributed equally to this work.

## Abstract

**Background:** Since December 2019, a cluster of coronavirus disease 2019 (COVID-19) occurred in Wuhan, Hubei Province, China and spread rapidly from China to other countries. In-hospital mortality are high in severe cases and cardiac injury characterized by elevated cardiac troponin are common among them. The mechanism of cardiac injury and the relationship between cardiac injury and in-hospital mortality remained unclear. Studies focused on cardiac injury in COVID-19 patients are scarce.

**Objectives:** To investigate the association between cardiac injury and in-hospital mortality of patients with confirmed or suspected COVID-19.

**Methods:** Demographic, clinical, treatment, and laboratory data of consecutive confirmed or suspected COVID-19 patients admitted in Wuhan No.1 Hospital from 25^th^ December, 2019 to 15^th^ February, 2020 were extracted from electronic medical records and were retrospectively reviewed and analyzed. Univariate and multivariate Cox regression analysis were used to explore the risk factors associated with in-hospital death.

**Results:** A total of 110 patients with confirmed (n=80) or suspected (n=30) COVID-19 were screened and 48 patients (female 31.3%, mean age 70.58±13.38 year old) among them with high-sensitivity cardiac troponin I (hs-cTnI) test within 48 hours after admission were included, of whom 17 (17/48, 35.4%) died in hospital while 31 (31/48, 64.6%) were discharged or transferred to other hospital. High-sensitivity cardiac troponin I was elevated in 13 (13/48, 27.1%) patents. Multivariate Cox regression analysis showed pulse oximetry of oxygen saturation (SpO_2_) on admission (HR 0.704, 95% CI 0.546-0.909, per 1% decrease, p=0.007), elevated hs-cTnI (HR 10.902, 95% 1.279-92.927, p=0.029) and elevated d-dimer (HR 1.103, 95%CI 1.034-1.176, per 1mg/L increase, p=0.003) on admission were independently associated with in-hospital mortality.

**Conclusions:** Cardiac injury defined by hs-cTnI elevation and elevated d-dimer on admission were risk factors for in-hospital death, while higher SpO_2_ could be seen as a protective factor, which could help clinicians to identify patients with adverse outcome at the early stage of COVID-19.

## Introduction

Since December 2019, a cluster of severe acute respiratory syndrome coronavirus 2 disease now named as 2019 novel coronavirus infection disease (COVID-19) occurred in Wuhan, Hubei Province, China and spread rapidly from China to other countries[1][2][3]. Cardiac injury and arrhythmias were common, especially for ICU patients [1][4]. Short-term prognosis of COVID-19 patients are discrepancy and in-hospital mortality risk are high in severe cases[1][2] Although previous study had indicated that several risk factors were independently associated with short-term mortality, such as elevated d-dimer, older age and higher Sequential Organ Failure Assessment (SOFA) score[2], few studies focused on cardiac injury with COVID-19 patients. High-sensitivity cardiac troponin I (hs-cTnI) was highly sensitive and specific markers of myocardial damage [5] and hs-cTnI elevation was not uncommon with COVID-19 patients[1][2]. However, a recent report of autopsy for a COVID-19 patient found that there were a few interstitial mononuclear inflammatory infiltrates, but no other substantial damage in the heart tissue [6]. Until now, the relationship between cardiac injury and in-hosptial prognosis and the mechanism of cardiac injury in COVID-19 patients remained controversial. Studies focused on cardiac injury in COVID-19 patients were scarce. In the present study, we sought to investigate the association between cardiac injury and in-hospital mortality of patients with COVID-19.

## Method

### Study design and participants

All consecutive patients with confirmed or suspected 2019 novel coronavirus infection disease (COVID-19) according to WHO interim guidance admitted in Wuhan No.1 Hospital from 25^th^ December 2019 to 15^th^ February 2020 were retrospectively screened. Wuhan No.1 Hospital is a reginal comprehensively central hospital which was responsible for the treatments for COVID-19 assigned by the government since 14^th^ February 2020. Before 14^th^ February 2020, patients with confirm or suspected COVID-19 were transferred to designated hospital after initial treatment. A portion of suspected patients with COVID-19 at the early stage of the outbreak did not receive severe acute respiratory syndrome coronavirus 2 (SARS-CoV-2) RT-PCR testing due to the limited supplies of diagnostic detection reagents. Patients who underwent hs-cTnI test within 48 hours after admission were included. Oral consents of all patients were obtained. This study was approved by the Wuhan No.1 Hospital institutional ethics committee. A comparison between non-survivors and survivor had been done and characteristics between patients who were included or not were also compared.

### Data collection

Demographic, clinical, treatment, laboratory data and in-hospital outcomes including death, transferred to designated hospital and discharge were extracted from electronic medical records.

### Laboratory procedures

Throat-swab specimens were obtained for SARS-CoV-2 detection from suspected patients using real-time PCR assay, which was performed by local Centers for Disease Control and Prevention and Wuhan No.1 Hospital. Routine blood examinations were complete blood count, serum biochemical tests (including renal and liver function, creatine kinase, lactate dehydrogenase, and electrolytes) were done for all inpatients on admission while the examinations of hs-cTnI, coagulation profile and C reactive protein were determined by the treating physician. Clinicians tended to perform hs-cTnI test on patients who were vulnerable to cardiac injury by their judgement.

### Definitions

Confirmed cases and suspected cases and the illness severity of COVID-19 was defined according to the Chinese management guideline for COVID-19 (version 5.0 or 6.0). The date of disease onset was defined as the day when the symptom was noticed. The durations from onset of disease to hospital admission were recorded. Fever was defined as axillary temperature of at least 37.3°C. The criteria for discharge were absence of fever for at least 3 days, substantial improvement in both lungs in chest CT, clinical remission of respiratory symptoms, and two throat-swab samples negative for SARS-CoV-2 RNA obtained at least 24 h apart. Cardiac injury was defined if the serum levels of high-sensitivity troponin I (hs-cTnI) were above the 99th percentile upper reference limit (0.026ug/L). Survivor group was composed of patients who was discharged or transferred to other hospital while non-survivor group included patients who died during hospital stay. Patients were followed up until they met survival endpoints.

### Statistical Analysis

Continuous variables with normal distribution were presented as mean±SD while were presented as median (IQR). Categorical variables were presented as n (%). We used the Student’s t test, Mann-Whitney U test, or Fisher’s exact test to compare differences between survivors and non-survivors where appropriate. To explore the risk factors associated with in-hospital death, univariate and multivariate Cox regression models were used. Considering the total number of deaths (n=17) in our study and to avoid overfitting in the model, five variables were chosen for multivariable analysis on the basis of previous findings, results of baseline characteristics comparison and clinical constraints. In a recent large cohort[2], Older age and d-dimer elevation was related to in-hospital death. SpO_2_ decrease was more common with severe patients than general patients. Chronic kidney diseases and elevated serum creatinine was more common among non-survivors [2]. Cardiovascular disease, lymphocytopenia and C reactive protein were excluded because their between-group differences were not significant. Severe type was excluded due to collinearity with SpO_2_. Between-group differences of leukocytosis, anemia, thrombocytopenia were statistically significant but we did not included them duo to unclear clinical significant. Hence, age, SpO_2_, serum creatinine value, d-dimer value analyzed as continuous variates and hs-cTnI elevation analyzed as categorical variate were chosen ultimately for our multivariable Cox regression model. Kaplan-Meier curve with log-rank test was used to compared mortality between patients with or without myocardial injury (MI). A two-sided α of less than 0.05 was considered statistically significant. Statistical analyses were done using the SPSS software (version 19).

## Result

From 25^th^ December 2019 to 15^th^ Feb 2020, a total of 110 patients(45.5% female, mean age 64.03±16.54 year old)with suspected (n=30, 27.3%) or confirmed (n=80, 72.7%) COVID-19 were admitted in department of respiration or emergency department of Wuhan No.1 Hospital. Cough was the most common onset symptom (n=38, 34.5%), followed by fever (n=33, 30.0%), fatigue (n=15, 13.6%), shortness of breath (n=10, 9.1%), lack of appetite (n=5, 4.5%), vomiting (n=3, 2.7%), dizziness (n=2, 1.8%), diarrhea (n=2, 1.8%), abdominal distention (n=1, 0.9%) and pharyngalgia (n=1, 0.9%). Median duration from symptom onset to admission was 7.0(IQR3.0-9.0) days. After median 9.0(IQR,6.0-12.0)days of hospitalization, death, discharge and transfer were 28 (25.5%), 64 (58.2%) and 18 (16.4%) patients respectively. A total of 64(58.2%)patients underwent hs-cTnI test during hospitalization. Among them, 48(75.0%, 48/64)patients who had their first hs-cTnI test within 48h after admission were included in our study. Among the remaining 16 patients, first hs-cTnI test performed between 4∼7 days, 8∼14 days and ≥14 days were in 7, 6 and 3 patients, respectively. All the other laboratory tests described in methods were done within 48h after admission. Inclusion flow chart was shown in Figure 1.

**Figure 1:**
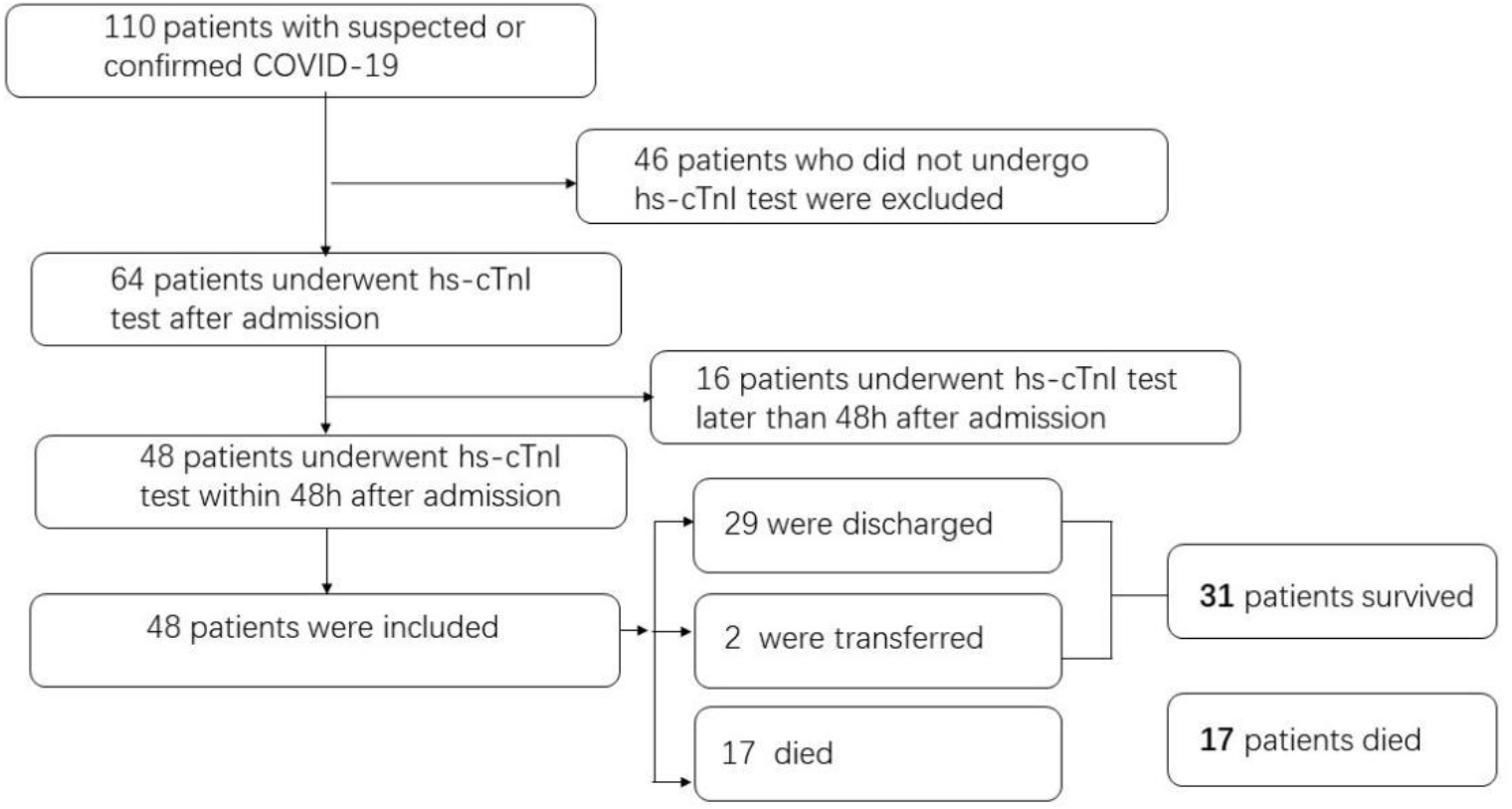
Inclusion flow chart.

A brief comparison was made between patients who was included (n=48) or not (n=62) in our study (TABLE 1) and found that patients included were older and male, hypertension, coronary artery disease, atrial fibrillation were more common among patients included. Disease severity status and treatment were similar. In-hospital mortality was higher in patients included (35.4% vs 17.7%, p=0.045).

**TABLE 1.** Clinical characteristics of patients included or excluded in the present study

|  | Total<br>(N=110) | Patients included<br>(n=48) | Patients excluded<br>(n=62) | *p-value* |
| --- | --- | --- | --- | --- |
| Age | 64.03±16.54 | 70.58±13.38 | 58.95±17.05 | **< 0.001** |
| Female | 50 (45.5) | 15 (31.3) | 35 (56.5) | **0.012** |
| Male | 60 (54.5) | 33 (68.8) | 27 (43.5) | **0.012** |
| Confirmed COVID-19 | 80 (72.7) |  |  | 0.281 |
| Disease severity status |  |  |  |  |
| General | 63 (57.3) | 26 (54.2) | 37 (59.7) | 0.698 |
| Severe | 47 (42.7) | 22 (45.8) | 25 (40.3) | 0.698 |
| Follow-up<br>(days) | 9.0 (6.0,12.0) | 10.0 (6.3,13.0) | 8.0 (5.0,12.0) | 0.099 |
| Comorbidity |  |  |  |  |
| HTN | 57 (51.8) | 32 (66.7) | 25 (40.3) | **0.007** |
| Diabetes | 19 (17.3) | 10 (20.8) | 9 (14.5) | 0.450 |
| CAD | 16 (14.5) | 13 (27.1) | 3 (4.8) | **0.002** |
| Previous stroke | 18 (16.4) | 11 (22.9) | 7 (11.3) | 0.123 |
| AF | 9 (8.2) | 7 (14.6) | 2 (3.2) | **0.040** |
| CKD | 9 (8.2) | 5 (10.4) | 4 (6.5) | 0.500 |
| Therapy |  |  |  |  |
| Antiviral | 108 (98.2) | 47 (97.9) | 61 (98.4) | > 0.999 |
| Antibiotic | 105 (95.5) | 46 (95.8) | 59 (95.2) | > 0.999 |
| Corticosteroids | 53 (48.2) | 22 (45.8) | 31 (50.0) | 0.704 |
| IVIG | 16 (14.5) | 6 (12.5) | 10 (16.1) | 0.786 |
| Oxygen inhalation | 71 (64.5) | 29 (60.4) | 42 (67.7) | 0.547 |
| NIMV | 37 (33.6) | 18 (37.5) | 19 (30.6) | 0.542 |
| IMV | 2 (1.8) | 1 (2.1) | 1 (1.6) | > 0.999 |
| In-hospital death | 28 (25.2) | 17 (35.4) | 11 (17.7) | 0.047 |

Note: Data are median (IQR), n (%), or n/N (%), where N is the total number of patients with available data. p values comparing patients included and excluded are from $\chi^2$ test, Fisher's exact test, or Mann-Whitney U test. HTN, Hypertension; CAD, cardiovascular disease; AF, atrial fibrillation; CKD, chronic kidney disease; IVIG, intravenous immunoglobulin; NIMV, non-invasive mechanical ventilation; IMV, invasive mechanical ventilation.

The mean age of 48 patients included was 70.58±13.38 year old and 15(31.3%)patients were female. After a median follow-up of 10 days, 17 patients (17/48, 35.4%) died while 29 (29/48, 60.4%) and 2 (2/48, 4.2%) were discharged or transferred to other hospital, respectively. Baseline clinical characteristics of 48 patients were shown in TABLE 2 compared by non-survivor and survivor. Patients died during hospital stay was older and severer on admission and the SpO_2_ was lower. Chronic kidney disease was more common among non-survivor while hypertension, coronary artery disease, diabetes, previous stroke and atrial fibrillation was similar between two gruops. The comparison of laboratory test shown higher value of serum creatinine, C reactive protein, d-dimer, white blood cell count but lower with hemoglobin and platelet in non-survivors. More intensive respiratory support was given to non-survivor due to the severity status but few patients were given invasive ventilation.

**TABLE 2.** Clinical characteristics of 48 patients on admission

|  | Total<br>(N=48) | Non-survivor<br>(n=17) | Survivor<br>(n=31) | *p-value* |
| --- | --- | --- | --- | --- |
| Age | 70.58±13.38 | 78.65±8.31 | 66.16±13.66 | **0.001** |
| Female | 15 (31.3) | 5 (29.4) | 10 (32.3) | > 0.999 |
| Confirmed COVID-19 | 32 (66.7) | 15 (88.2) | 17 (54.8) | **0.026** |
| Disease severity status |  |  |  |  |
| General | 26 (54.2) | 4 (23.5) | 22 (71.0) | **0.002** |
| Severe | 22 (45.8) | 13 (76.5) | 9 (29.0) | **0.002** |
| Time from onset to admission (days) | 7.0 (3.0,10.0) | 7.0 (3.0,11.5) | 7.0 (3.0,9.0) | 0.354 |
| Fever | 34 (70.8) | 12 (70.6) | 22 (71.0) | > 0.999 |
| Follow-up | 10.0 (6.3,13.0) | 12.0 (7.0,17.0) | 9.0 (6.0,12.0) | 0.153 |
| (days) |  |  |  |  |
| Comorbidity |  |  |  |  |
| HTN | 32 (66.7) | 12 (70.6) | 20 (64.5) | 0.757 |
| Diabetes | 10 (20.8) | 5 (29.4) | 5 (16.1) | 0.295 |
| CAD | 13 (27.1) | 4 (23.5) | 9 (29.0) | 0.747 |
| Previous stroke | 11 (22.9) | 6 (35.3) | 5 (16.1) | 0.163 |
| AF | 7 (14.6) | 3 (17.6) | 4 (12.9) | 0.686 |
| CKD | 5 (10.4) | 5 (29.4) | 0 (0.0) | **0.004** |
| HR beats per min | 80.0 (74.0,98.0) | 88.0 (78.5,104.0) | 80.0 (74.0,92.0) | 0.113 |
| RR breaths per min | 20.0 (19.0,21.8) | 20.0 (19.0,24.0) | 20.0 (19.0,20.0) | 0.207 |
| SBP (mmHg) | 129.19±14.41 | 130.29±16.67 | 128.58±13.27 | 0.698 |
| DBP (mmHg) | 77.67±12.94 | 77.06±14.34 | 78.00±12.34 | 0.812 |
| SpO<sub>2</sub> (%) | 93.27±4.95 | 91.12±5.36 | 94.45±4.35 | **0.024** |
| SpO<sub>2</sub>% < 90% | 13 (27.1) | 7 (41.2) | 6 (19.4) | 0.173 |
| SpO<sub>2</sub>% < 95% | 26 (54.2) | 11 (64.7) | 15 (48.4) | 0.368 |
| hs-cTnI (ug/L) | 0.012<br>(0.005,0.031) | 0.034<br>(0.011,0.247) | 0.006<br>(0.002,0.014) | **0.001** |
| hs-cTnI ≥0.026ug/L | 13 (27.1) | 10 (58.8) | 3 (9.7) | **<0.001** |
| CK (U/L) | 85.0<br>(52.0,197.0) | 104.5<br>(50.5,293.5) | 83.0<br>(60.0,158.0) | 0.508 |
| CKMB (U/L) | 9.0 (7.0,14.0) | 9.5 (7.3,21.3) | 9.0 (7.0,11.0) | 0.206 |
| ALT (U/L) | 21.0 (12.5,34.0) | 19.0 (11.0,31.0) | 23.0 (15.0,39.0) | 0.257 |
| AST (U/L) | 32.0 (22.0,51.8) | 33.0 (19.0,67.0) | 32.0 (22.0,49.0) | 0.738 |
| LDH (U/L) | 293.0<br>(217.0,546.0) | 302.0<br>(193.0,1009.0) | 291.0<br>(219.0,415.0) | 0.745 |
| Serum Cr (umol/L) | 80.0 (66.3,97.5) | 96.0 (76.0,184.0) | 79.0 (64.0,90.0) | **0.014** |
| CRP (mg/L) | 43.2 (18.5,107.9) | 65.5 (46.4,125.9) | 23.8 (15.3,87.6) | **0.006<sup>‡</sup>** |
| Fbg (mg/L) | 4.57±1.47 | 4.40±1.48 | 4.66±1.49 | 0.574<sup>§</sup> |
| D-Dimer (mg/L) | 0.625 (0.323,2.035) | 2.295<br>(0.413,17.005) | 0.565<br>(0.313,0.943) | **0.025<sup>§</sup>** |
| WBC (×10<sup>9</sup>/L) | 6.485 (4.145,8.825) | 8.84 (5.46,9.67) | 5.74 (4.14,7.39) | **0.017** |
| Leukocytosis<br>( > 9.5×10<sup>9</sup>/L) | 5 (10.4) | 4 (23.5) | 1 (3.2) | **0.047** |
| Lymphocyte (×10<sup>9</sup>/L) | 0.985 (0.620,1.308) | 0.92 (0.51,1.29) | 1.08 (0.66,1.33) | 0.223 |
| Lymphocytopenia<br>( < 1.1×10<sup>9</sup>/L) | 26 (54.2) | 10 (58.8) | 16 (51.6) | 0.765 |
| Hb (g/L) | 122.92±21.02 | 111.35±23.96 | 129.26±16.40 | **0.004** |
| Anemia ( < 110g/L) | 13 (27.1) | 9 (52.9) | 4 (12.9) | **0.006** |
| PLT (×10<sup>9</sup>/L) | 167.0<br>(116.0,217.5) | 119.0 (88.5,212.5) | 181.0<br>(146.0,220.0) | **0.047** |
| Thrombocytopenia<br>( < 125×10<sup>9</sup>/L) | 13 (27.1) | 9 (52.9) | 4 (12.9) | **0.006** |
| Therapy |  |  |  |  |
| Antiviral | 47(97.9) | 17(100) | 30 (96.8) | > 0.999 |
| Antibiotic | 46 (95.8) | 17 (100) | 29 (93.5) | 0.533 |
| Corticosteroids | 22 (45.8) | 8 (47.1) | 14 (45.2) | > 0.999 |
| IVIG | 6 (12.5) | 4 (23.5) | 2 (6.5) | 0.167 |
| Oxygen inhalation | 29 (60.4) | 2 (11.8) | 27 (87.1) | **< 0.001** |
| NIMV | 18 (37.5) | 14 (82.4) | 4 (12.9) | **< 0.001** |
| IMV | 1 (2.1) | 1 (5.9) | 0 (0.0) | 0.354 |

Note: Data are median (IQR), n (%), or n/N (%). p values comparing patients included and excluded are from $\chi^2$ test, Fisher's exact test, or Mann-Whitney U test. HR, heart rate; RR, respiratory rate; SBP, systolic blood pressure; DBP, diastolic blood pressure; SpO<sub>2</sub>, Pulse Oxygen Saturation; Fbg, fibrinogen; Hb, hemoglobin. a: non-survivor(n=17) vs survivor(n=27); b: non-survivor(n=16) vs survivor(n=29); c: non-survivor(n=14) vs survivor(n=26).

High sensitivity-cTnI elevated in 13(27.1%)patients while 35(72.9%)patients had hs-cTnI valve within normal range. After median 10.0(IQR, 6.3-13.0)days of hospitalization, 17 (35.4%) patients (10 patients in hs-cTnI group and 7 patients in normal gourp) died and 31 (64.6%) were discharged or transferred to designated hospital. Mortality was significantly higher among patients with hs-cTnI elevation(76.9% vs 20.0%, P<0.001)(Figure 2). Kaplan-Meier curve shown that in-hospital mortality were higher among patients with elevated hs-cTnI (log-rank p=0.005)(Figure 3). Multivariate Cox regression analysis shown that pulse oximetry of oxygen saturation (SpO_2_) on admission (HR 0.704, 95% CI 0.546-0.909, per 1% decrease, p=0.007), elevated hs-cTnI (HR 10.902, 95% 1.279-92.927, p=0.029) and elevated d-dimer (HR 1.103, 95%CI 1.034-1.176, per 1mg/L increase, p=0.003) on admission were independently associated with in-hospital mortality (TABLE 3).

**TABLE 3.** Univariate and multivariate Cox regression analysis

| Variates | Univariate |  |  | Multivariate |  |  |
| --- | --- | --- | --- | --- | --- | --- |
|  | HR | 95% CI | p-value | HR | 95% CI | p-value |
| Age | 1.029 | 0.981-1.079 | 0.245 | 1.008 | 0.937-1.085 | 0.825 |
| SpO<sub>2</sub>% | 0.876 | 0.769-0.997 | 0.046 | 0.704 | 0.546-0.909 | **0.007** |
| Serum Cr value | 1.003 | 1.001-1.004 | 0.002 | 1.002 | 0.999-1.005 | 0.118 |
| d-dimer value | 1.072 | 1.028-1.118 | 0.001 | 1.103 | 1.034-1.176 | **0.003** |
| hs-cTnI elevation | 3.588 | 1.335-9.641 | 0.011 | 10.902 | 1.279-92.927 | **0.029** |

Note: Cox proportional hazard regression analysis was conducted to investigate the effects of variates on predicting in-hospital mortality. HR, hazard ratio; CI, confidence interval.

**Figure 2.**
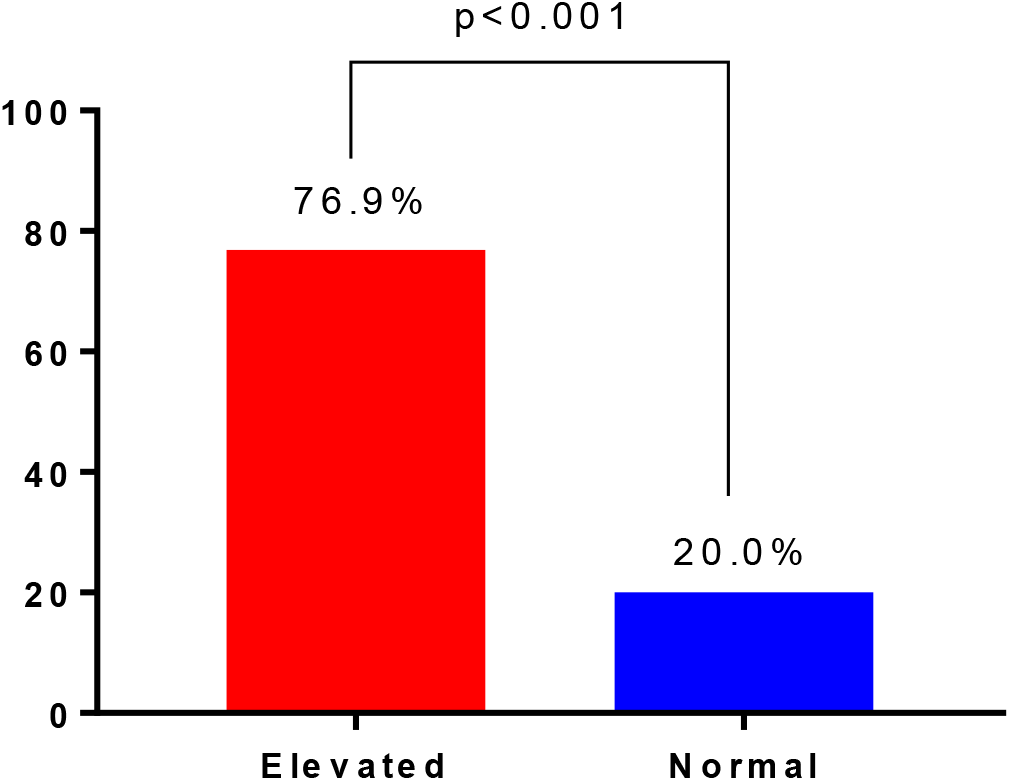
Mortality between patients with or without elevated hs-cTnI.

**Figure 3.**
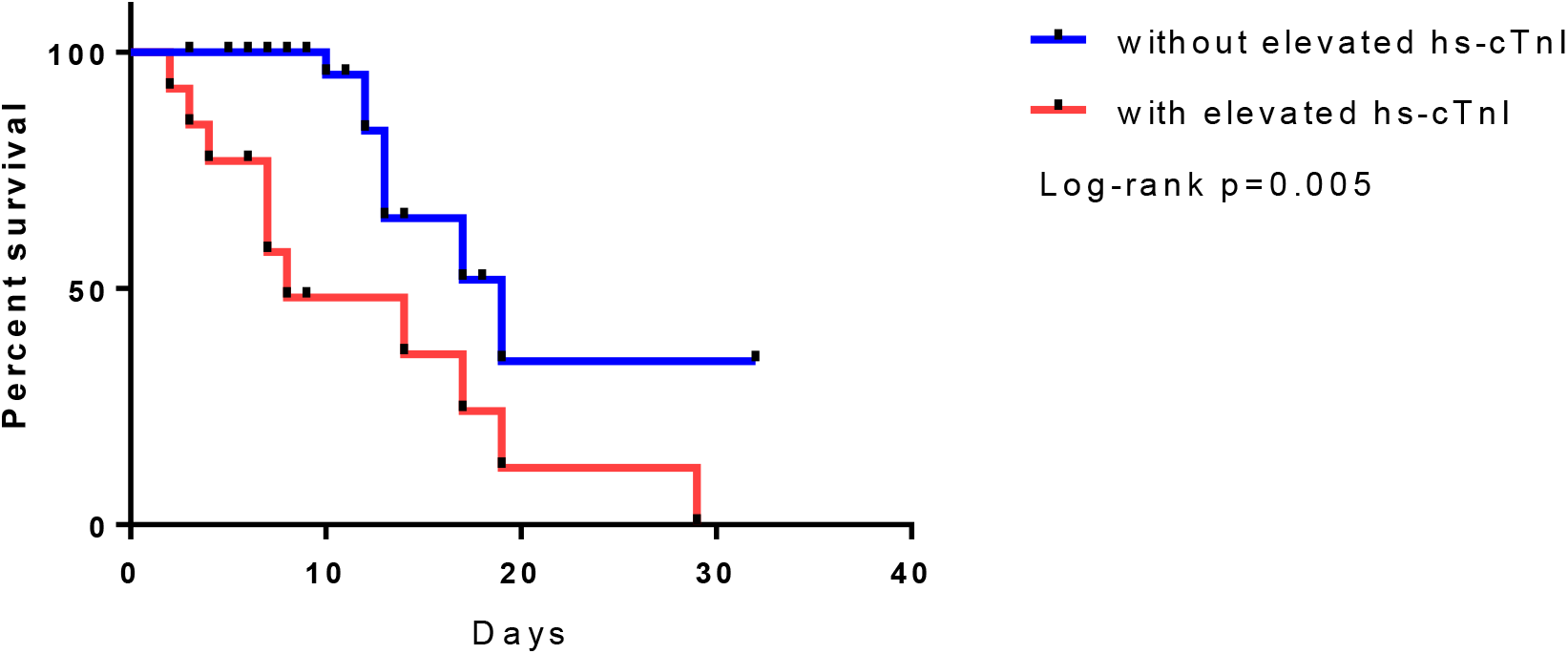
Kaplan-Meier Curve.

## Discussion

Obviously there is an major concern globally regarding the emerging outbreak of COVID-19, with increasing cases confirmed in multiple countries currently. The World Health Organization has upgraded its coronavirus risk assessment to “very high” at global level as COVID-19 continuously spread in more countries despite rigorous global containment and quarantine efforts. Recent studies concerning the novel coronavirus provided evidence of the potential connection SARS-CoV-2 infection and acute cardiac injury[2, 4]. We observed that the clinical symptoms of COVID-19 and other viral pneumonia were highly variable and overlap, whereas the most common symptoms were cough, fever and fatigue, with dizziness and diarrhea less common. While typical symptoms like fever could be absent in the disease progression particularly in elderly, adversely, gastrointestinal discomfort and altered mental status presented more frequently. High sensitivity cTnI tests were more inclined to perform on participants with older age and preexisting concomitants include hypertension, cardiovascular disease and atrial fibrillation.

Mortality rate was higher significantly in the older individuals, might explained by decaying organ systems,particularly respiratory tract, immune system and cardiovascular system and urinary system. In addition, more remarkable laboratory abnormalities was observed within severe patients and those with poor outcomes. The concentration of CRP was considerably higher among the deceased than the survived ones. The average hemoglobin count was obviously lower than the survivors and over half deaths sustained with anemia,suggesting a relatively status of hypoxia and malnutrition. The occurrence of leukocytosis might accompany with a secondary bacterial infection which has higher frequency in non-survivors.

Thrombocytopenia was revealed in more than half non-survivors whereas not to the degree of depletion, which provided extra evidence that the coagulation was highly activated. We analyzed the coagulation marker as manifested by d-dimer, notably revealing that there were substantial coagulation abnormalities in COVID-19 patients, in addition, more profoundly among severe and fatal illness. Most non-survivors underwent oxygen administration,which we unfortunately discovered whether noninvasive nor invasive mechanical ventilation could reverse the rapidly deterioration of multi-systemic damage. The multivariate Cox regression model showed that elevation of hs-cInI and the levels of SpO2% and d-dimer were independent predictors of in-hospital prognosis of COVID-19. Evidently, the elevation of hs-cT and D-dimer and decreased SpO2 was associated with higher risk of death.

Current reports suggest that majority of patients on admission for pneumonia underwent coagulation disorders, which included higher platelet aggregating activity, elevated levels of antithrombin and thrombin–antithrombin complex, decreased factor IX activity[7, 8]. The association between SARS-CoV-2 infection and acute myocardial injury has been appreciated, likewise stronger when the infection is more severe. Limited reports of autopsy detected cardiomyocytes necrosis and degeneration,with intercellular substance infiltrated with small amounts of monocytes, lymphocyte and/or neutrophils, additionally, the occurrence of endoangiitis and thrombosis was noted [9]. No evidence has shown that SARS-CoV-2 infected myocardium directly, despite the putative receptor of SARS-CoV-2, ACE2 [10], same with SARS-CoV, as well as the documented reports state that the RNA of SARS-CoV has been identified within myocardium of affected patients [11]. While confirmation of SARS-CoV-2-related myocarditis has been hampered by its heterogeneous clinical presentations and complexity of definitive diagnose, PCR technology have enabled detection of many other virus within the myocardium of affected patients [12]. Hence, whether the virus exerted its pathogenic effects by transition to myocardium directly is yet to investigated. Apart from that, it’s likely the host response to infection is major determinants in COVID-19 patients that are complicated by myocardial injury [13].

It’s estimated that pneumonia can affect the cardiovascular system by multiple mechanisms. The established pneumonia associated with SARS-CoV-2 accounted for the inciting infection, results in relative or severe hypoxemic state of the host, prompted by impaired gas exchange across the alveoli of inflamed lung parenchyma and ventilation-perfusion mismatching. Moreover, inflammation increase the metabolic needs of peripheral tissues and organs, attributable to myocardial ischemia, may related either to: increasing heart rate which shortens the diastolic duration, coronary artery spasm or coronary microvascular dysfunction, without ruptured plaque within coronary atherosclerosis, therefore compromising coronary perfusion and exaggerating cardiac metabolic imbalance [7]. The preexistent cardiovascular diseases and COVID-19 appears to accelerate the progression bidirectional. On the other hand, circulating inflammatory factor and toxin mediated vasoconstriction adversely affect myocardial function,through which called cytokine storm. The overall effect of the inflammatory response triggered by infection is dictated by the balance between pro-inflammatory (eg, IL-1, IL-6, TNF-α, IL-8) and anti-inflammatory (eg, IL-10,TGF-β) mediators [14]. Sustained or uncontrolled release of cytokines secreted by various cell populations especially immune cells, inflicting excessive systemic inflammation which leads to multiple organ dysfunction including lung and heart [15]. In this case, we can suggest both viral and following bacterial infections hijack the defense mechanism of host cells by targeting immune cell-mediated cytokine signaling and contributes to the cytokine storm.

It’s fully anticipated that respiratory infections and sepsis are associated with artery and venous thrombosis along with microvascular embolism. Cardiac thrombosis could be generated locally induced by either exposure to tissue factors following vascular damage,or spontaneously under procoagulant condition secondary to an exaggerated inflammation which stimulated endothelial and mononuclear cells, eventually provoking the coagulation cascade. In addition to clotting activation, systemic coagulation abnormalities also involves suppression of fibrinolysis [8, 16, 17]. Dehydration, elderly, obesity, basic diseases, long-term bedridden, clinical practice of glucocorticoid and immunoglobulin,invasive operation multifactorial,as previously summarized, were the main risk factors of hyperviscosity, which intensified the coagulation disorder. Our data suggest that the enhanced procoagulant activity was inherent with ACI incidence among COVID-19 inpatients. Awareness of this information should guide clinical intervention aimed at pro-coagulation disturbance in high-risk groups.

## Limitations

There were several limitations in our study. A substantial proportion of suspected cases was included and patients without hs-cTnI test was excluded, both of which might resulted in some bias. Also, our results based on a very small cohort. The data of hs-cTnI test before symptom onset was unavailable and we could not tell the acute cardiac injury form the chronic cardiac injury. Electrocardiography and echocardiography was not evaluated in our study. A part of patients were transferred to other hospital and no definite outcomes could be evaluate. But we used Cox regression analysis to solve this limitation.

## Conclusion

Cardiac injury defined by hs-cTnI elevation and elevated d-dimer on admission were risk factors for in-hospital death, while higher SpO_2_ could be seen as a protective factor, which could help clinicians to identify patients with adverse outcome at the early stage of COVID-19.

## Data Availability

All data presented in this manuscript are from the Wuhan No.1 Hospital. And these data have already approven to publish. There are no patient privacy or medical ethics problems.

## Notes

### Competing Interest Statement

The authors have declared no competing interest.

### Clinical Trial

This is a retrospective study.

### Funding Statement

No external funding was received.

